# Shut it down: a cross country panel analysis on the efficacy of lockdown measures

**DOI:** 10.1101/2020.04.12.20062695

**Authors:** Vincenzo Alfano, Salvatore Ercolano

**Affiliations:** Department of Structures for Engineering and Architectures, University of Napoli Federico II; Department of Mathematics, Computer Science, and Economics, University of Basilicata

**Keywords:** COVID-19, coronavirus, Lockdown, Panel data, Government measures

## Abstract

Coronavirus pandemic outbreak from China in the December 2019 and since then has quickly spread all over the world. National governments introduced policies aimed to reduce the probability to contract the virus, such as lockdown measures, in order to limit the outbreak. Lockdown fostered a debate about the effective need and the optimal duration of such measures. Indeed, these policies have a high price, being characterized by the alt of many productive activities. The aim of this note is to provide preliminary evidences about the efficacy of lockdown measures all over the world, by the means of a panel data quantitative analysis. Our results confirm the efficacy of such measures, and that the average time to have effects in terms of a reduction of cases is of about ten days. Furthermore the beneficial effects of a lockdown keep reducing the new cases with a linear trend for at least the ten successive days.

## 1. Introduction and research question

Diffusion of COronaVIrus infectious Disease (COVID-19) began in China, during December 2019, when the first cases were identified in the province of Wuhan. Since that moment the COVID-19 has quickly spread all over the world. For this reason, on March the 11^th^ 2020, the World Health Organization (WHO) declared COVID-19 a pandemic. According to WHO data on April the 1st 2020 there were more than 1.5 millions of confirmed cases, about 93,000 confirmed deaths and at least 212 countries, areas or territories reported confirmed cases of infection. USA are observing a fast growth of COVID-19 and today it is the first country for confirmed cases (with about 478,000 cases). Europe also presents a massive concentration of COVID-19, with more than 803,000 confirmed cases. Spain is today the most damaged European country, with over 157,000 confirmed cases; it is followed by Italy (147,577), Germany (119,624) and France (117,749).

Even if the approach followed by national governments in order to face the emergency has been very heterogenous across countries (Piguillem and Shi, 2020), it is possible to identify two main kind of policies: i) health policies aimed to strengthen the capacity of the hospital system to deal with the effects of COVID-19 (such as plans for expansion of health workforce, support to companies producing medical supplies, and so on); ii) policies aimed to reduce the probability to contract the virus (such as lockdowns and social distancing measures). In this work we focus on the latter family of policy, and namely on lockdowns: nevertheless it is important to notice that provisions belonging to both these two families of provisions are deeply interconnected. As a matter of fact, while health policies directly support the national health systems, lockdown and social distancing aim to ease the strain on health services by slowing down the outbreak (Hamzelou, 2020). According to Shao (2020), “adding a large number of hospital beds along with city lockdown improved cure rates and reduced mortality rates. Residential unit lockdown effectively control the spread of the epidemic, and sooner implementation of this measure resulted in more effective epidemic control”. Summing up a very complex and heterogeneous framework, it is possible to detect that a combination of containment and mitigation activities has been carried out by national governments, pursuing the objective to delay surges of patients, contain the demand for hospital beds and protect the most exposed population (Bedford et al, 2020).

This research focuses on lockdown, since this policy has fostered a debate about the effective need of such measures. It is likely that this resistance in the stakeholders and the policymakers is due to the fact that lockdown comes with a very important economic price, being characterized by the necessary alt of many productive activities. In fact, despite the fact that both WHO and previous literature focused on the Chinese case (De Figureido et al 2020; Lau et al, 2020) highlights the importance and centrality of such measures in order to reduce the probability of contagion (and thus the related diffusion of the virus), the political debate appeared to be very influenced by the negative impact of such a measure on national economies. In this rationale, the debate has been focused on the potential inefficiency of this policy, or also on the estimation of a good trade-off between safeguarding citizens’ health and keeping healthy the national economy. To give some examples of such a debate, the web site www.eurotopics.net^1^collected several declarations about the risk of lockdown: two former leading Czech bankers declared to “have serious doubts that the risks of the coronavirus are indeed so great that they justify intervention without discussion and time limits and the use of such instruments. Yes, people expect the government to provide security and protect their lives. But certainly not at any cost^2^.”. Further anecdotical evidence includes declarations on this line on prominent newspapers from Ireland^3^, Netherlands^4^, but also countries stroke more heavily from the pandemic, such as Italy^5^ and Germany^6^.

In order to clear the public debate from such perspectives, it is very important to study how lockdown effectively works, how effective it is, and to empirically test the efficiency of such a measure in terms of reducing the contagion. Different contributions are trying to address this important issue, from different perspectives. Nevertheless to the very best of our knowledge, the main empirical contributions specifically focused on the efficacy of lockdown have been carried out at national level, often adopting SIR models, and especially focusing on the Indian and on the Chinese case. It is important to notice that given the novelty of the topic, many of this work are still working papers. These literature includes Lau et al. (2020), that conclude that due to lockdown measures, a significantly decreased growth was observed in China; and Sardar et al. (2020), that using a predictive analytical study that incorporates the lockdown measures for India, conclude that the positive effects of the lockdown can be observed just in some provinces. Piguillem and Shi (2020) study the topic from a theoretical perspective, adapting a SIR model to include lockdown and testing measure: their theoretical finding is that lockdown is a second best option to the testing. On the other hand, moving to a cross country perspective, the main contributions carried out comparisons among the different patterns followed by the virus (Kévorkian et al. 2020) but to the best of our knowledge at the moment no empirical evidence on the impact of lockdown measures is provided. Cruz and Dias (2020), using a qualitative approach, investigated COVID-19 in four countries (China, Italy, Brazil and USA) and suggest that “not all relevant actions were taken, in a timely manner, to efficiently address the spread of Covid-19”.

The aim of this note is to provide the first empirical evidence about the efficacy of lockdown measures by the means of a quantitative analysis through a panel data approach. In more details, gathering data on the diffusion of the virus from the John Hopkins University dataset and on Government measures from ACAPS, we try to address two different research questions, namely:

Q1) From a cross country perspective, is there an empirical evidence about the capability of lockdown measures to reduce the number of contagions?

Q2) After how longs lockdown measures became effective? How their efficiency change over time?

We consider both of this points to be very important in order to provide some evidences about the efficacy and the related timing of such policies.

The present short note is organized as follow: in the next section we briefly introduce the data and methodology employed in the present analysis, in section three we report the main results and some preliminary conclusions are discussed in the last section.

## 2. Data and methods

The aim of this research is to empirically estimate the impact of lockdown measures on the spread of the virus. It is indeed suggested by the literature (Piguillem and Liyan, 2020) that the lockdown is one of the most effective policies to prevent the spread of the pandemic; nevertheless surprisingly to the best of our knowledge the economic empirical literature has not yet payed very much attention to the theme. This is the first estimation attempt assuming a cross-country perspective. To pursue this objective, we utilizes a panel dataset, with daily data from several countries from all over the world used as the basic statistical unit of observation. In formal terms, we are going to estimate the following equation:

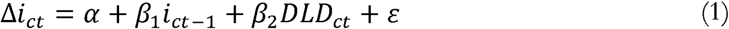

Where Δ*i* is the amount of new COVID-19 cases at time *t* (with respect to time *t*-1) in country *c*; it is modelled as a function of the infected in the country *c* during the previous day (*i*_*t*−1_), which of course is an important variable in determine the number of new infected given the very nature of a pandemic; furthermore the equation includes a dichotomous dummy DLD, that signals if in the day *t* there was a lockdown in act in country *c*, or not; alternatively, DLD may signal which country had a lockdown in place since *x* days (more details on this later on), in order to control per the time efficiency of the policy. As usual, identifies the error term.

In order to estimate our equation, two kinds of information are needed: i) the daily amount of COVID-19 cases and ii) the measures of lockdown put in place. In order to obtain the first kind of data we relied as a source on the *Novel Coronavirus Cases* dataset, compiled by the Johns Hopkins University Center for Systems Science and Engineering (JHU CCSE) from various sources,^7^ in its last version available while writing this research (which is the one of April the 11 ^th^). It offers daily estimation of the COVID-19 cases since January the 22^nd^ up to April the 10^th^ for 211 countries (oversea territories of some European countries, such as for instance French Martinique or Netherlands Antilles, and more in general out-lands territories are considered different countries to these purposes, given the difficulty of spreading the virus within this far away administrative units, even if technically belonging to the same country; please also consider that only 200 of these countries are included in our final sample, given the listwise deletion of some minor insular Pacific countries due to lack of data per the other variables datasets). This offers us the best possible data on the impact and the dynamic of the COVID within a country. From this source is computed our dependent variable *New cases*, the operationalization of Δ*i*, as the first difference between the cases of today and the one of yesterday, and also one of the dependent variable, *YCases*, the operationalization of *i*_*t*−1_, which is the absolute value of cases found the day before.

For the second source of information our dataset relies on the ACAPS data gathered from the COVID-19: Government Measures Dataset. This let us to distinguish between countries that applied lockdown measures, and countries that did not. More specifically, we used the last version available at the time of the writing of this article (the one of April the 9^th^) and build the dichotomous dummy variable *Dummy Lockdown* (operationalization of DLD) that assumes the value of 1 in the first date that a country implemented a partial or complete lockdown measure with respect to the entire population (we obtain the set of countries and dates imposing three filters, namely: *Introduction/extension of measures* as log type; *Lockdown* as category; and No as target) and for all the subsequent days in which the lockdown is held. The choice of including only policies aimed to the entire population is justified in order to avoid having biased estimation due to policy intervention that were referred to a small share of the population. This strategy lead to a result of a total of 100 measures captured by the DLD dummy, in 67 different countries, in many different dates (from January the 4^th^ per Mauritius and Nepal, to April the 9^th^ per Armenia). Furthermore we also computed different DLD with respect to the days elapsed since the implementation of the policy. As a matter of fact, 97.5% of those who develop symptoms do it within 11.5 days from the infection, which means a confidence interval from 8.2 to 15.6 days (Lauer et al., 2020), or in other terms being in place a certain delay between the actual infection and the possibility of being tested positive, the lockdown may have greater benefits in terms of a reduction of the new cases after a certain time since the imposition of the policy is elapsed. Descriptive statistics about the variables are presented in table 1.

**Table 1.**
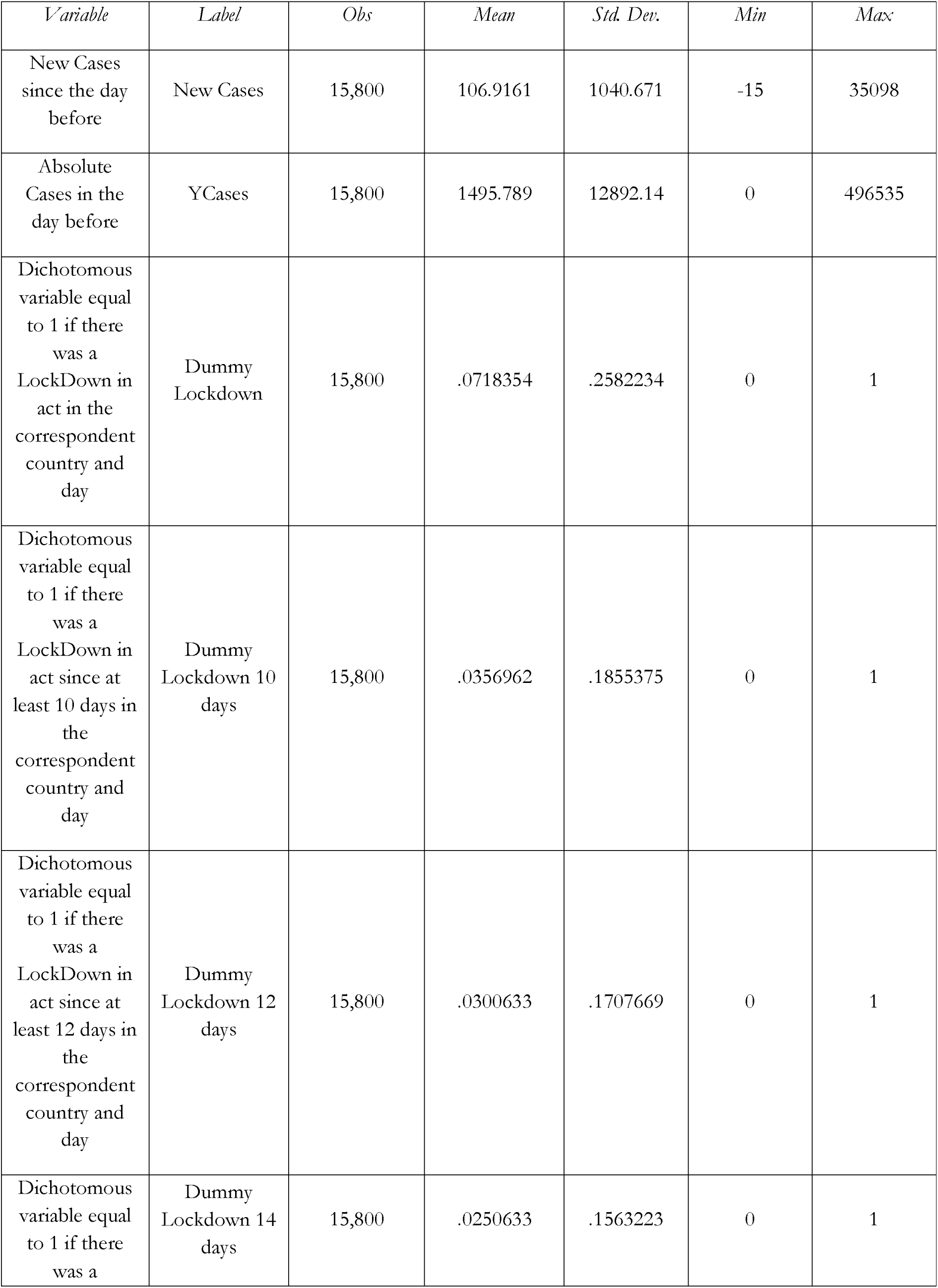
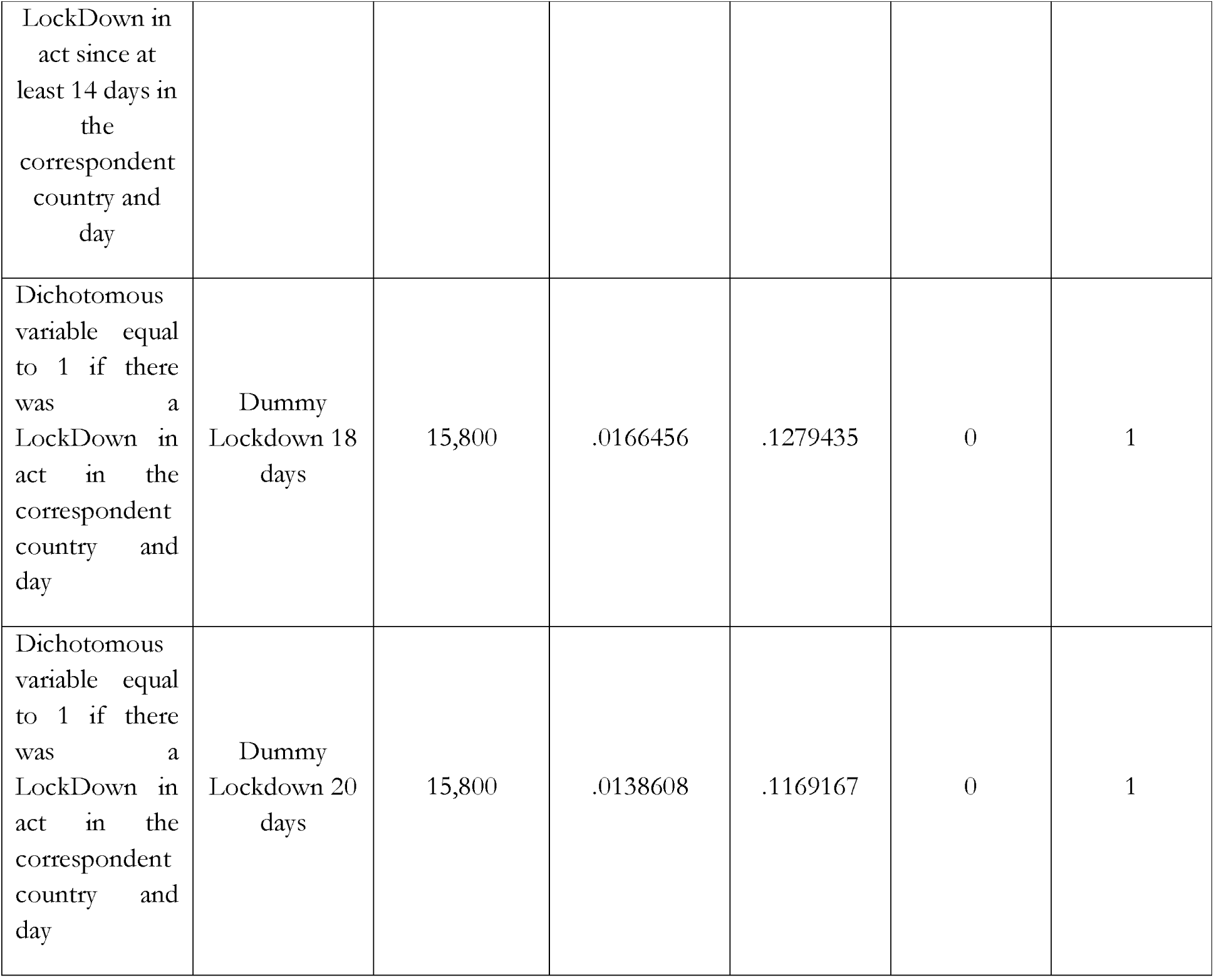
Descriptive statistics

Considering the panel nature of our dataset, that has several observation per each *c* and *t*, the best estimator to compute the impact of DLD is a Feasible – General Least Square (F-GLS), that corrects the coefficient per the repeated nature of the date (Aigner and Balestra, 1988; Hsiao, 1986). Indeed, given the large *t* in our data (79 different periods) estimators from the General Method of Moments family do not seem an ideal choice (Arellano and Bond, 1991).

Moreover, considering that the spread of the virus may be due several factors specific to each country, such as its geographical location, its proximity to other countries infected, its climate, its demography and so on and so forth, we consider appropriate use an estimator, such as the F-GLS with fixed effects, that captures the heterogeneity between countries. In other words, our aim is to estimate the effects with respect to the single countries, assuming that the heterogeneity among them does not change in the 80 days of our time span. For all this reasons, it is appropriate to use an estimation technique that helps us clearing our coefficients from all the possible biases due to this spurious relationships or omitted variables. Indeed, as a matter of fact, with a fixed effect estimation we measure the dynamic within each and any country.

The final dataset is composed by 79 daily observation (per 80 days, since January the 22^th^ to April the 9^th^; this is given the loss of one observation per computing the new cases) per 200 countries, for a total of 15,800 observation in the most complete specification. Other than world-wide, we also computed separated continental estimates, under the assumption that local political and geographical characteristic may play a role into this relationship. For this reason, it is relevant that of these 200 countries, 47 are in Europe (and thus a total of 3,555 observations are included in this subsample); 45 are in Asia (for a total of 3,713 observations); 54 are in Africa (for a total of 4,266 observations) and 48 in America (for a total of 3,792 observations). The remaining six countries are in the Pacific region (Australia, Fiji, French Polynesia, New Caledonia, New Zealand and Papua New Guinea).

## 3. Results

Results of the estimates through a F-GLS estimator with Fix Effects on the whole sample are provided in table 2. As it is possible to see, *YCases* has a positive coefficient and is statistically very significant in all the different specifications, confirming of course that the dependent variable *New Cases* mainly depends on the amount of cases of the day before.

**Table 2.** F-GLS Regression - World

|  | (2.1) | (2.2) | (2.3) | (2.4) | (2.5) | (2.6) |
| --- | --- | --- | --- | --- | --- | --- |
|  | New cases | New cases | New cases | New cases | New cases | New cases |
| YCases | 0.0751<sup>***</sup> | 0.0757<sup>***</sup> | 0.0758<sup>***</sup> | 0.0757<sup>***</sup> | 0.0754<sup>***</sup> | 0.0754<sup>***</sup> |
|  | (183.70) | (185.29) | (185.93) | (186.24) | (186.48) | (187.59) |
| Dummy<br>Lockdown | -147.1<sup>***</sup> |  |  |  |  |  |
|  | (-7.77) |  |  |  |  |  |
| Dummy<br>Lockdown 10<br>days |  | -381.4<sup>***</sup> |  |  |  |  |
|  |  | (-13.77) |  |  |  |  |
| Dummy<br>Lockdown 12<br>days |  |  | -467.8<sup>***</sup> |  |  |  |
|  |  |  | (-15.11) |  |  |  |
| Dummy<br>Lockdown 14<br>days |  |  |  | -542.2<sup>***</sup> |  |  |
|  |  |  |  | (-15.36) |  |  |
| Dummy<br>Lockdown 18<br>days |  |  |  |  | -709.4<sup>***</sup> |  |
|  |  |  |  |  | (-13.64) |  |
| Dummy<br>Lockdown 20<br>days |  |  |  |  |  | -1042.9<sup>***</sup> |
|  |  |  |  |  |  | (-15.99) |
| Constant | 13.21<sup>***</sup> | 15.43<sup>***</sup> | 15.76<sup>***</sup> | 15.33<sup>***</sup> | 14.06<sup>***</sup> | 16.69<sup>***</sup> |
|  | (3.11) | (3.73) | (3.82) | (3.72) | (3.41) | (4.05) |
| Observations | 15800 | 15800 | 15800 | 15800 | 15800 | 15800 |

t statistics in parentheses
\* $p < 0.1$ , \*\* $p < 0.05$ , \*\*\* $p < 0.01$

The *Dummy Lockdown* has a negative and statistically very significant coefficient, suggesting that in the world on average countries that have implemented the lockdown have less New Cases that country who did not. While looking at the several Dummies Lockdown, that signal the presence of a lockdown since respectively at least 10, 12, 14, 18 or 20 days, we can see that there are still benefits, and that the benefits of the lockdown in terms of less infected increases linearly with the passing or the days.

While looking at the European countries subsample (table 3), the situation slightly differs. Europe has been hardly hit by the COVID-19, and relatively to other continents it is densely inhabited and with several different states that exist in a relatively small space. Even in that case the YCases variable is positive and statistically significant, suggesting the relationship between the New Cases and the cases of yesterday still holds. The Dummy Lockdown is positive, suggesting that in countries that implemented the lockdown there are on average more New Cases than in countries who did not. This is possibly due to the fact that in Europe, lockdown measures have been implemented on average when the diffusion of the virus was quickly growing. Moreover in some of the countries that implemented the lockdown, such as Italy and Spain, the dynamic of COVID-19 spread is ahead compared to other countries that chose to do not impose an early lockdown (possibly refusing to take advantage of the information about the outbreak in neighbourhood countries). In that rationale, makes sense that this variable has a positive coefficient, since it just signals the presence in the same continent of countries at different stage of the spreading of the virus, or in other terms at different distances from the peak of the infection curve, and the relative different choices of the governments. Nevertheless it is important to notice that also in Europe this difference becomes statistically non-significant while comparing countries that implemented the lockdown since at least 10 days, with the ones who did not. Furthermore, while comparing countries who implemented the lockdown since at least 12 days with the rest of the sample, the coefficient becomes negative, suggesting a net benefit of this policy after 12 days; its magnitude and statistical significance grows after 14, 18 and 20, and does it with a more than proportional (i.e. exponential) trend.

**Table 3.** F-GLS Regression - Europe

|  | (3.1) | (3.2) | (3.3) | (3.4) | (3.5) | (3.6) |
| --- | --- | --- | --- | --- | --- | --- |
|  | New cases | New cases | New cases | New cases | New cases | New cases |
| YCases | 0.0492<sup>***</sup> | 0.0542<sup>***</sup> | 0.0563<sup>***</sup> | 0.0570<sup>***</sup> | 0.0566<sup>***</sup> | 0.0577<sup>***</sup> |
|  | (51.34) | (52.26) | (54.41) | (56.40) | (61.23) | (65.30) |
| Dummy<br>Lockdown | 532.7<sup>***</sup> |  |  |  |  |  |
|  | (11.74) |  |  |  |  |  |
| Dummy<br>Lockdown 10<br>days |  | 97.75 |  |  |  |  |
|  |  | (1.39) |  |  |  |  |
| Dummy<br>Lockdown 12<br>days |  |  | -149.2<sup>*</sup> |  |  |  |
|  |  |  | (-1.90) |  |  |  |
| Dummy<br>Lockdown 14<br>days |  |  |  | -291.6<sup>***</sup> |  |  |
|  |  |  |  | (-3.34) |  |  |
| Dummy<br>Lockdown 18<br>days |  |  |  |  | -412.8<sup>***</sup> |  |
|  |  |  |  |  | (-3.66) |  |
| Dummy<br>Lockdown 20<br>days |  |  |  |  |  | -1060.2<sup>***</sup> |
|  |  |  |  |  |  | (-8.22) |
| Constant | 55.40<sup>***</sup> | 81.96<sup>***</sup> | 84.14<sup>***</sup> | 84.11<sup>***</sup> | 83.38<sup>***</sup> | 83.53<sup>***</sup> |
|  | (5.47) | (8.15) | (8.38) | (8.40) | (8.33) | (8.41) |
| Observations | 3713 | 3713 | 3713 | 3713 | 3713 | 3713 |

t statistics in parentheses
\* $p < 0.1$ , \*\* $p < 0.05$ , \*\*\* $p < 0.01$

## 4. Conclusions

This short note aimed to provide some first empirical evidences in a cross country perspective about the efficacy and efficiency of lockdown measures in terms of reducing the contagions. Using a quantitative approach on panel data, our preliminary results seem to confirm that this policy has a positive impact in term of reduction of COVID-19 cases. We also pointed out some specificity between the estimations carried out considering countries all over the world and European countries, which are likely to be due to a difference in the concentration of the virus.

Nevertheless our results seem to confirm the efficacy of lockdown and its exponential effects over time. These results could contribute to shed some light in a growing debate about the effective need of such policy, due to the trade-off in place between the reduction of the diffusion of the disease and the negative impact lockdown has on economic activities. It is also worth noting that while these are preliminary results, that need further analysis, certainly could contribute to the academic and political debate about the definition of “mitigation measures” and the possible “exit strategy” from lockdown. For instance while so far the “exit strategy” proposed is the one by Sarwal and Sarwal (2020), that suggest a “localization” strategy after 14 days of lockdown, which according to the authors represent a sort of deadline where no further gains can be expected by extending the lockdown, our results seem to contradict this finding, and detect an increase in the benefit and a growing reduction of contagions also 20 days after the lockdown. It is nevertheless important to highlight also in this conclusion that our perspective is the one of a cross-country analysis, and thus average for the world or a continent, with all the benefits in terms of generalization of the results, but also the limits while applying this finding to a specific country or to derive a precise estimate. Caution is suggested in reading this results, that are certainly also driven by the timing of the measures in Europe and the rest of the world, other than the spreading of the pandemic. For this reason we feel obliged to highlight the importance and the need of further investigations on this topic.

## Data Availability

All data are described and public available

## Footnotes

1 https://www.eurotopics.net/en/237330/what-are-the-risks-of-lockdown, url consulted on April the 10^th^ 2020.

2 Hospodářské noviny, 20 March, 2020

3 The Irish Times fears: “People won’t put up with months of lockdown.”The Imperial College report is less ambiguous: social distancing needs to be in force for at least two-thirds of the time until a vaccine becomes available. That’s 18 months away. The risk, during that period, is that resentment will fester. Right now, we are frightened and grateful for strong and stable leadership, but fatigue and irritation will set in. People who are bored and anxious are susceptible to voices offering other, more appealing scenarios and solutions, or those who say there was another way. Down the road, a whole other crisis of democracy is looming.” The Irish Times 18 March, 2020

4 De Volkskrant pointed out that there is no proof of the effectiveness of the lockdown policy: “In the short term, a lockdown might have some effect, but it would not prevent more severe corona outbreaks in the long term. Neither the expected increase in corona deaths in the Netherlands nor the fact that other European countries are ordering a stricter - but equally unchecked - regime are reasons for a change of course. Of course it’s conceivable that in an unpredictable crisis like this the government might change its course of action, but this must be based on new insights from the experts, not on the vague assumption that the more radical interventions are in principle the better ones.” De Volkskrant, 19 March, 2020

5 Political scientist Nadia Urbinati complained on HuffPost Italia: “We are being given to understand that the entire responsibility lies with the citizens. But what about the responsibility of the institutions which today are threatening to take even ‘tougher’ measures? Is there amnesia about the decisions taken in the recent past - those that have trampled on and weakened public Health? The decisions of governments at both the national and regional level must be taken into account when allocating responsibility. Yet we have not heard a word of self-criticism.” Huffpost Italy, 18 March, 2020

6 Contemporary historian René Schlott also had an uneasy feeling about the drastic shutting down of public life. In a guest commentary for the Süddeutsche Zeitung he writes: “If we didn’t know better we could interpret the events of recent days for a right-wing populist takeover. However once a precedent has been set, who can rule out the possibility that the same restrictions on fundamental rights will be reactivated again in the future in the name of another supposed emergency? The closure of the border with Austria, partly on the grounds that it prevents panic buying from the neighbouring country, is a slap in the face to all those who, until a few days ago, defended the policy of open borders. The fundamental right to asylum has been made obsolete. The humanitarian dam has burst.” Süddeutsche Zeitung, 17 March, 2020

7 Namely it is compiled with data from: the World Health Organization (WHO), DXY.cn. Pneumonia. 2020, BNO News, National Health Commission of the People’s Republic of China (NHC), China CDC (CCDC), Hong Kong Department of Health, Macau Government, Taiwan CDC, US CDC, Government of Canada, Australia Government Department of Health, European Centre for Disease Prevention and Control (ECDC), Ministry of Health Singapore (MOH).

## References

Aigner, D.J. and Balestra, M. (1988). Optimal Experimental Design for Error Components Models, Econometrica, vol. 56, No. 4, (July 1988) 955–971.

Arellano, M. and Bond, S. (1991). Some tests of specification for panel data: Monte Carlo evidence and an application to employment equations. Review of Economic Studies. 58 (2): 277.

Bedford, J., Enria, D., Giesecke, J., Heymann, D. L., Ihekweazu, C., Kobinger, G., & Ungchusak, K.(2020). COVID-19: towards controlling of a pandemic. The Lancet.

Cruz, B. S., & de Oliveira Dias, M. (2020). COVID-19: From Outbreak to Pandemic. GSJ, 8 (3).

Hsiao, C. (1986). Analysis of Panel Data, Cambridge University Press, Cambridge.

Kevorkian, A., Grenet, T., & Gallee, H. (2020). Covid-19 pandemic: on a simple way to visualize the epidemic states and trajectories of some European countries, and to assess the effect of delays in official response. medRxiv.

Figueiredo A M, Daponte Codina A, Figueiredo M., Saez M & Cabrera León A. Impact of lockdown on COVID-19 incidence and mortality in China: an interrupted time series study. [Submitted]. Bull World Health Organ. E-pub: 6 April 2020. doi: http://dx.doi.org/10.2471/BLT.20.256701

Hamzelou, J. (2020). World in lockdown.New Scientist Volume 245, Issue 3275

Lau, H., Khosrawipour, V., Kocbach, P., Mikolajczyk, A., Schubert, J., Bania, J., & Khosrawipour, T. (2020). The positive impact of lockdown in Wuhan on containing the COVID-19 outbreak in China. Journal of Travel Medicine.

Lauer SA, Grantz KH, Bi Q, et al. (2020). The Incubation Period of Coronavirus Disease 2019 (COVID-19) From Publicly Reported Confirmed Cases: Estimation and Application. Ann Intern Med. 2020;

Piguillem, F., & Shi, L. (2020).The optimal covid-19 quarantine and testing policies (No. 2004). Einaudi Institute for Economics and Finance (EIEF).

Sardar, T., Nadim, S. S., & Chattopadhyay, J. (2020). Assessment of 21 Days Lockdown Effect in Some States and Overall India: A Predictive Mathematical Study on COVID-19 Outbreak. arXiv preprint 2004.03487.

Sarwal, R., & Sarwal, T. (2020). Mitigating COVID-19 With Lockdowns: A Possible Exit Strategy. Available at SSRN 3563538.

Shao, P. (2020). Impact of city and residential unit lockdowns on prevention and control of COVID-19. medRxiv.

